# Identification of CAA as highly specific and sensitive antibody target for acute schistosomiasis diagnostics

**DOI:** 10.1101/2023.03.07.23286891

**Authors:** Anna O. Kildemoes, Tom Veldhuizen, Stan T. Hilt, Lisette van Lieshout, Taniawati Supali, Erliyani Sartono, Maria Yazdanbakhsh, Daniel Camprubí-Ferrer, Jose Muñoz, Joannes Clerinx, Paul L.A.M. Corstjens, Govert J. van Dam, Leo G. Visser, Meta Roestenberg, Angela van Diepen, Cornelis H. Hokke

## Abstract

**Background:** The WHO 2030 roadmap for schistosomiasis calls for development of highly sensitive and specific diagnostic tools to continue and sustain progress towards elimination. Serological assays are excellent for sensitive detection of primary schistosome infections and for schistosomiasis surveillance in near- and post-elimination settings. To develop accurate assay formats, it is necessary to identify defined antibody targets with low cross-reactivity and potential for standardized production. Here we focus on defined schistosome glycan antigens.

**Methods:** Target identification was performed by assessing antibody responses in well characterized cross-sectional and cohort sample sets (n=366 individuals) on tailor-made antigen microarrays. IgM and IgG binding to candidate diagnostic targets was measured for serum/plasma samples from: controlled human schistosome infection model (CSI), schistosome infected travellers, soil-transmitted helminth infected and non-infected individuals.

**Findings:** We found that antibodies to circulating anodic antigen (CAA) identify schistosome infection with high sensitivity (IgM≥100%, IgG≥97%) and specificity (IgM≥93%, IgG≥97%) in the test samples. Infection dose affected timing of anti-CAA antibody isotype switch. Furthermore, we demonstrate the presence of shared and non-specific glycan epitopes in crude schistosome cercarial and egg antigen preparations. Many non-specific glycan epitopes contained in crude antigen mixes are responsible for a large proportion of false schistosomiasis positives in standard serological assays.

**Interpretation:** CAA is target for development of highly sensitive and specific tools for schistosomiasis serology with use cases for travellers and surveillance in near and post-elimination settings as well as emerging transmission zones.

**Funding:** Global Health Innovative Technology Fund (GHIT), HIC-Vac, Leiden University Medical Center (LUMC)

## INTRODUCTION

Schistosomiasis caused by parasitic blood flukes remains a large global public health burden with an estimated cost of 1.9 million disability-adjusted life years (DALYs) and more than 250 million people infected worldwide (1). The current WHO 2021-2030 roadmap for schistosomiasis control and elimination advocates for holistic and integrated strategies to tackle the disease burden.

Development of diagnostics is identified as a critical need to accelerate progress towards elimination (2). For non-endemic, low endemic, and near- or post-elimination settings, highly specific diagnostic tools for surveillance are needed. Additionally, strategies combining sensitive first line screening tests with second line assays specific for active infections are envisioned to optimize resource use and drug impact (3). Serological assays are excellent for sensitive detection of primary infections with schistosomes and are attractive for feasible transmission surveillance. High sensitivity is an inherent property of robust serological methods, which basically exploit the host driven signal amplification induced after antigen recognition and subsequent antibody response. However, a well-known limitation of diagnostic serology is the persistence of antibodies in circulation after successful clearance or cure of an infection. As antibody detection cannot distinguish adequately between prior and current infections, it adds the most value in the context of primary exposures such as in non-endemic area travel medicine and surveillance in emerging transmission risk zones. In endemic regions, serology can be a powerful element in transmission risk mapping as well as near- and post-elimination monitoring, particularly through strategic application in defined target groups such a young children or occupational risk groups (4, 5).

Typical antibody detection platforms used for schistosomiasis diagnosis are enzyme-linked immunosorbent assays (ELISA), indirect haemagluttinin assays (IHA) and/or immune fluorescence assays (IFA, commonly using adult worm sections). Many of such existing commercial and/or in house serological assays are based on crude soluble egg, cercarial or worm antigen preparations, which are undefined mixes of antigens and hence suffer from batch-to-batch variation as well as contain cross-reactive protein and glycan epitopes (6-8). In schistosome infection, a large proportion of the antibodies elicited recognise glycan elements present in the parasite’s glycoprotein and glycolipid repertoire, as demonstrated both in rodent and primate models as well as cross-sectional human sample sets (9-11). Subsets of glycan elements expressed in schistosomes are shared with other helminths, other invertebrates, food stuff of plant origin or the mammalian host (12-14). Anti-glycan antibodies that recognise glycans shared between schistosomes and other sources are an inherent liability to the specificity of assays based on crude antigen preparations. From a diagnostic perspective, it is therefore essential to identify which glycan targets are specific and which are not.

Extensive glycomics work confirms that schistosomes express several glycan elements that appear unique to the parasite, in addition to elements that also occur in other organisms (15). Here we exploit the antigenic properties of such unique glycan structures using controlled human schistosome infection (CSI) and primary infection traveller samples to identify targets for future development of diagnostic tools. Custom microarrays of a panel of crude, purified and synthetic antigens were constructed based on extensive life-stage stratified glycomics data of *S. mansoni* combined with published glycan repertoire data from other helminth species (15), and published anti-glycan antibody data from primate models and human sample sets (10, 11, 16). The repetitive polysaccharide structures circulating cathodic antigen (CCA) and circulating anodic antigen (CAA) abundant in the schistosome gut were also included. Gut-localised antigens are of particular interest as the reactivity seen in schistosomiasis IFAs is to the intestinal tissue on the adult worm sections (17). Specifically, the aim of this study is use these custom microarrays to identify defined glycan antigen(s) which display high specificity (≥95%) and sensitivity (≥95%) for development of highly accurate antibody-detection tools aimed at primary schistosome infection diagnosis.

## METHODS

### Anonymised serum sample sets

Additional sample and ethical clearance information in Suppl. Section 1

### Primary schistosome infection sample sets

A. Controlled human Schistosome Infection (CSI; n=17)) samples from 17 healthy adult Dutch volunteers infected with male-only *S. mansoni* cercariae (no egg production) (17).
B. Post-Travel Screening of Parasites (PTSP; n = 131 pre-travel, n=113 paired pre-post travel) study samples including nine cases diagnosed post-travel as schistosomiasis positive by seroconversion (18).
C. Spanish tourist cluster (n=6) exposed/infected with *S. mansoni-*clade parasites in Mozambique. All individuals were egg negative and presented with acute schistosomiasis symptoms (19).
D. Belgian tourist cluster (n=34) exposed/infected with *S. haematobium x S*.*matheii* parasites. All individuals were egg negative and 32/34 presented with acute schistosomiasis symptoms (20, 21).

### Schistosome infection-negative sample sets

E) Soil-transmitted helminth infection (STH) positive (*Ascaris* sp., *Trichuris trichiura, Necator americanus, Ancylostoma duodenale, Strongyloides stercoralis*) samples from Indonesia (n=97) (22).
F) *Strongyloides stercoralis* infection positive sera (n=25) from the LUMC biobank.
G) Sera obtained via the Dutch blood donor bank, Sanquin, from healthy blood donors (n=56).

### Construction of microarrays

Crude schistosome soluble cercarial antigen and egg antigen from both *S. mansoni* (SmCA, SmSEA) and *S. haematobium* (ShCA, ShSEA) was produced by mechanical disruption of parasite material in cold PBS followed by sonification (Branson Sonic Power Company, Sonifier B-12) and centrifugation (17000g). The material was split for sodium meta-periodate (NaIO_4_) and mock treatment for glycan epitope disruption. Mock treatment and NaIO_4_ treatment was done in pre-cooled 0.4M acetate buffer (pH 4.5, NaAc3H2O, JTBaker, 0256) with and without 40mM of NaIO_4_ (Merck, 1.0659) respectively (o/n cold incubation on roller). Quenching was done with one volume of 50mM sodium borohydride (NaBO_4_, Fluka, 71321) for 30 minutes on ice. All material was dialysed against 1x PBS o/n (Thermo-Scientific, Slide-a-Lyzer mini dialysis devices 3500MWCO). Synthetic glycan elements, keyhole limpet haemocyanin (Sigma-Aldrich, H7017), native schistosome circulating cathodic and anodic antigens (CCA, CAA), printbuffer controls, and crude schistosome antigen preparations with and without NaIO_4_ treatment were printed in 10%DMSO (Sigma-Aldrich D8418) in printbuffer (Nexterion Spot 1066029) in triplicates/array onto epoxilane coated glass slides (Nexterion Slide E, Schott 1066643) with a MicroGrid robot (BioRobotics) (23). Full target list and details can be found in supplementary table 1.

### Microarray incubations

Microarrays with fitted silicone gaskets were reconstituted in 1xPBS and blocked for a minimum of one hour (PBS, 2% bovine serum albumin (Sigma-Aldrich A3059) with 50mM ethanolamine (>99.5%, Sigma-Aldrich 411000)) on shaker. Each array was washed in PBS-Tween0,05%, PBS and 1:100 serum sample in buffer/array added for 1hour incubation at room temperature while shaking. For 8 sample gaskets a total of 250ul diluted sample/array was used whereas 30ul/array was used for 64 sample gaskets. Arrays were then washed (as above) and 1:1000 detection antibodies added (goat anti-human IgG-Cy-3, Sigma-Aldrich C2571; goat anti-human IgM-AF647, Invitrogen, A21249) for 30min room temperature incubation on shaker. For IgG subclass experiments 1:400 mouse anti-human (Southern Biotech) IgG1 Hinge-AF555 (#9052-32), IgG2 Fc-AF555 (#9070-32), and IgG3 Hinge-AF647 (#9210-31) were used and IgG4 pFc’-AF647 (#9190-31) in 1:250. Finally, slides were washed as above with an additional wash in milli-Q before they were spun dry and scanned (Agilent Scan Control™ 2006).

### Schistosomula culture and UCP-LF CAA and CCA measurements

*S. mansoni* cercariae were transformed by heat-shock. Briefly cercariae were shed from *Biomphalaria glabrata* snails in Bar-Le Duc water and then kept on ice for ∼two hours. Prewarmed culture media (Hybridoma-SFM (Gibco ref. 12045-076), Penicillin-streptomycin (Gibco ref. 15140122), 200 µM ascorbic acid (J.T. Baker ref. 1018), 1:500 chemically defined lipid concentrate (Gibco ref. 11905031)) was added to sedimented cercariae. Incubation was done in 37°C waterbath with mechanical mixing every five minutes for 20 minutes. Schistosomulae were separated from tails on orbital shaker and transferred for 20 minutes, 37°C incubation with antibiotics and antimycotic (1:100 ABAM (Sigma-Aldrich ref. A5955) in Dulbecco’s PBS (DPBS, Sigma-Aldrich D8662). Schistosomulae were sedimented and media changed to 10ml media with 0,02% red blood cells (Sanquin, washed in RPMI1640 HEPES, Gibco ref. 13018015) added for plating into 48 well plate (200µl parasites plus 800µl media/well).

Single schistosomulae baseline and day 8 samples were collected. For day 1-7 five full well replicates and a control were collected. Parasites were counted in 100µl per well at 2x magnification. The remaining material was split into supernatant and parasite material by centrifugation at 2000g.

Parasite pellets were washed twice in DPBS and subsequently freeze-dried, resuspended in DPBS and sonified. CAA and CCA were measured in supernatant and schistosomulae by UCP-LF BCAAhT17 and UCP-LF BCCAhT17 wet assays, respectively (urine assay protocols (24) with urine to PBS adaptation and 2% final concentration TCA). Cut-offs that apply to these culture media experiments are: UCP-LF BCAAhT17 = 10pg/ml, UCP-LF BCCAhT17 = 1000pg/ml (24, 25).

### Data processing and analyses

Microarray scan images were obtained through Agilent Feature Extraction 10.7.3.1 and imported into GenePixPro vs7. Spot morphology quality was inspected and median fluorescence intensity (MFI) values obtained per nanodot with the average of each triplicate target used for analyses. Further data processing was done in Microsoft Excel and graphs were generated using GraphPad Prism 9.3.1.

Variation is shown as standard deviations with horizontal bars indicating median values. Response profiles and magnitude of response to single targets can be compared between samples, but different targets within arrays are not directly quantitively comparable due to differences in printing characteristics determining spot density and epitope accessibility. An arbitrary theoretical cut-off value of MFI=10000 is used as comparative basis for specificity and sensitivity calculations for different antigen(s). Welch’s unequal variances t-test was used to compare mean CAA/CCA content in individually collected schistosomulae. Comparison of CAA/CCA content and excretion in the multi-well culture experiment was done using Brown-Forsythe and Welch’s ANOVA for with Dunnett’s T3 multiple comparisons test (GraphPad Prism 9.3.1).

## RESULTS

We first conducted a diagnostic candidate target down-selection based on a subset of controlled schistosome infection (CSI) samples. This time course sample set is ideal to identify accurate infection driven antibody responses that are low at baseline (selecting for high specificity) and robustly mounted by all individuals after infection (selecting for high sensitivity). Particularly, CSI samples are suitable to determine specificity cut-offs that are challenging to obtain from cross-sectional or cohort studies in endemic areas due to persistence of antibodies from prior infections. IgM and IgG binding to targets printed on microarrays were evaluated in CSI samples at baseline and 12/16 weeks post infection (infection dose = 20 cercariae, n=8 (table 1, individuals shown as I-VIII)). Criteria for selecting candidates of interest were low baseline signal (MFI<10000) and positive response at week 12 and/or 16 post infection (MFI>15000). Responses between MFI 10000 and 15000 were permitted in both categories for the first down-selection and a single non-complier per category was also allowed. This fluidity in cut-off was considered for first evaluation to avoid prematurely excluding targets of interest based on a small sample size. Targets were only considered relevant if they lived up to both specificity and sensitivity cut-off for IgM and/or IgG. Candidates fulfilling criteria for IgM and/or IgG are shown in bold and with thick border (table 1) and target details can be found in suppl. table 1. For IgM, four targets fulfilled the criteria in the first evaluation: *S. mansoni* soluble egg antigen (SmSEA) and CAA performed with 100% specificity and sensitivity, while di-LeX-HSA and di-Lex-BSA showed 87,5% specificity and 100% sensitivity. For IgG, six targets fulfilled the criteria: SmSEA-mock, SmSEA, di-CAA-BSA and FLDNFagal with specificity of 100% and sensitivity of 85,7%, *S. mansoni* cercarial antigen (SmCA)-per with specificity of 85,7% and sensitivity of 100%, while native purified CAA performed the best with 100% for both measures. A second evaluation was done on smaller arrays to reproduce and confirm observations in extended time-course sample series and increased sample size (CSI n=17). In addition to the synthetic di-LeX element, native CCA was included as this structure is known to consist of repetitive Lewis-X (LeX) motifs (26). The di-CAA element was not included in the second arrays as it is part of the repetitive structure of native purified CAA antigen (27). FLDNFagal, an abundant motif in the cercarial glycocalyx (30) which met the performance criteria in the first down-selection failed with respect to specificity upon assessing more CSI samples (IgM 70,6% (12/17); IgG 58.8% (10/17)) and is therefore not discussed in the following.

**Table 1:**
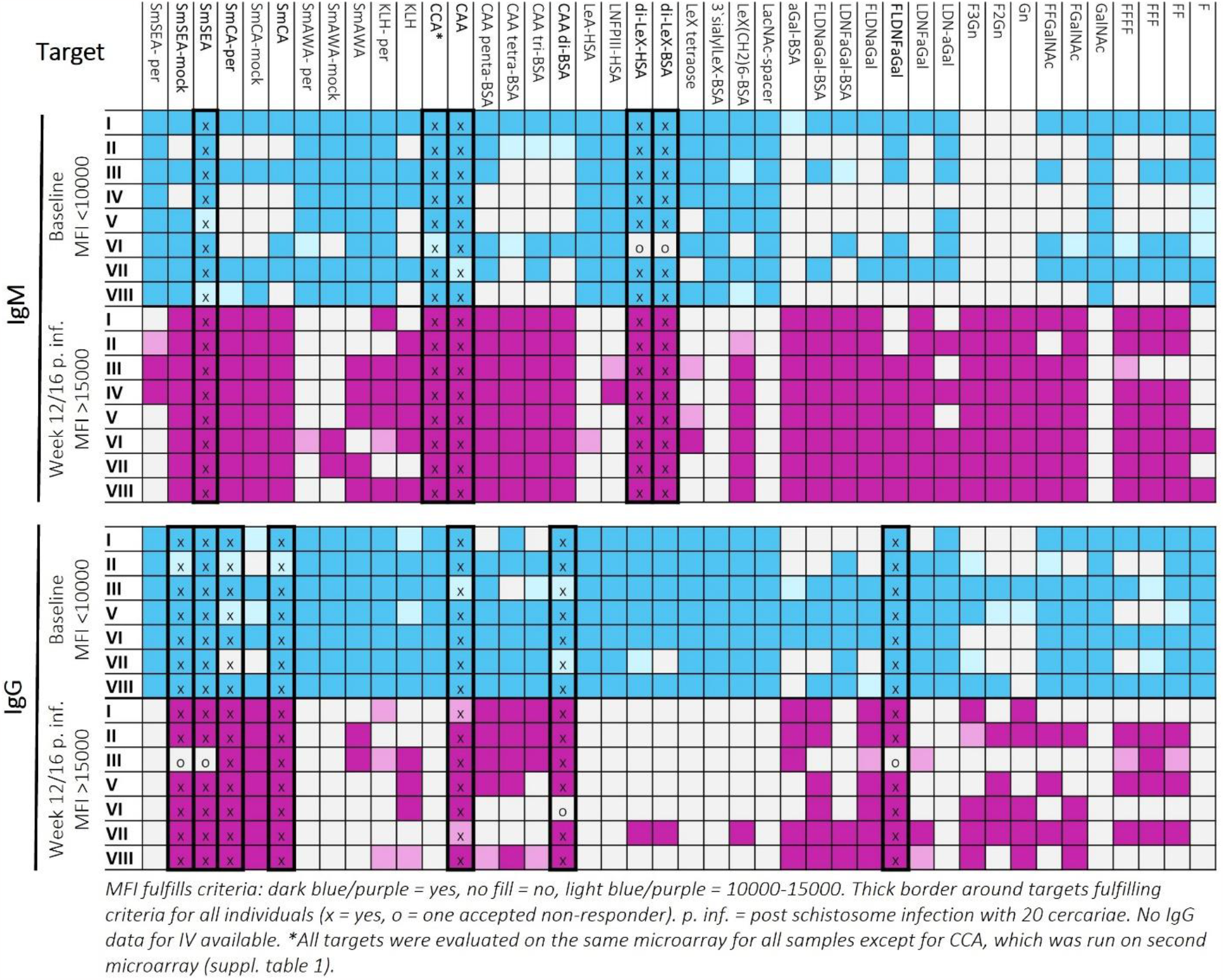
First microarray-based IgM and/or IgG target down-selection by individual level evaluation of antibody responses in controlled human infection samples (I-VIII)

### Glycan epitopes shared between schistosomes and other organisms are present in crude antigen

Time-course IgM and IgG responses to crude SmCA and SmSEA mixes from baseline to one year post infection in CSI participants present clear infection related antibody dynamics (Figure 1A, B). Albeit with variable background reactivity level at baseline. IgM responses are initiated uniformly upon infection for SmCA and SmSEA with average seroconversion at week three to four weeks post infection (arbitrary cut-off MFI=10000). Larger variation for the averaged IgG response curve is explained by more variation in onset of isotype switch from IgM to IgG on individual level (Figure 1B). The IgG subclass repertoire elicited in response to infection does not change from early (week four to eight) to later stage (week 12-20) of infection but does vary on inter-individual level in terms of which subclasses are elicited (Figure 2C,D). For example, individual IX has both IgG1 and IgG2 whereas X and XI predominantly induce IgG2. Comparison of periodate treated SmCA and SmSEA with their mock treated counter parts demonstrates that for both SmCA and SmSEA most antibody binding is to glycans. Periodate treatment disrupts monosaccharide ring structures with adjacent cis-hydroxyl groups, hence mediating a molecular change that abolish antibody binding to the majority of glycan epitopes.

**Figure 1:**
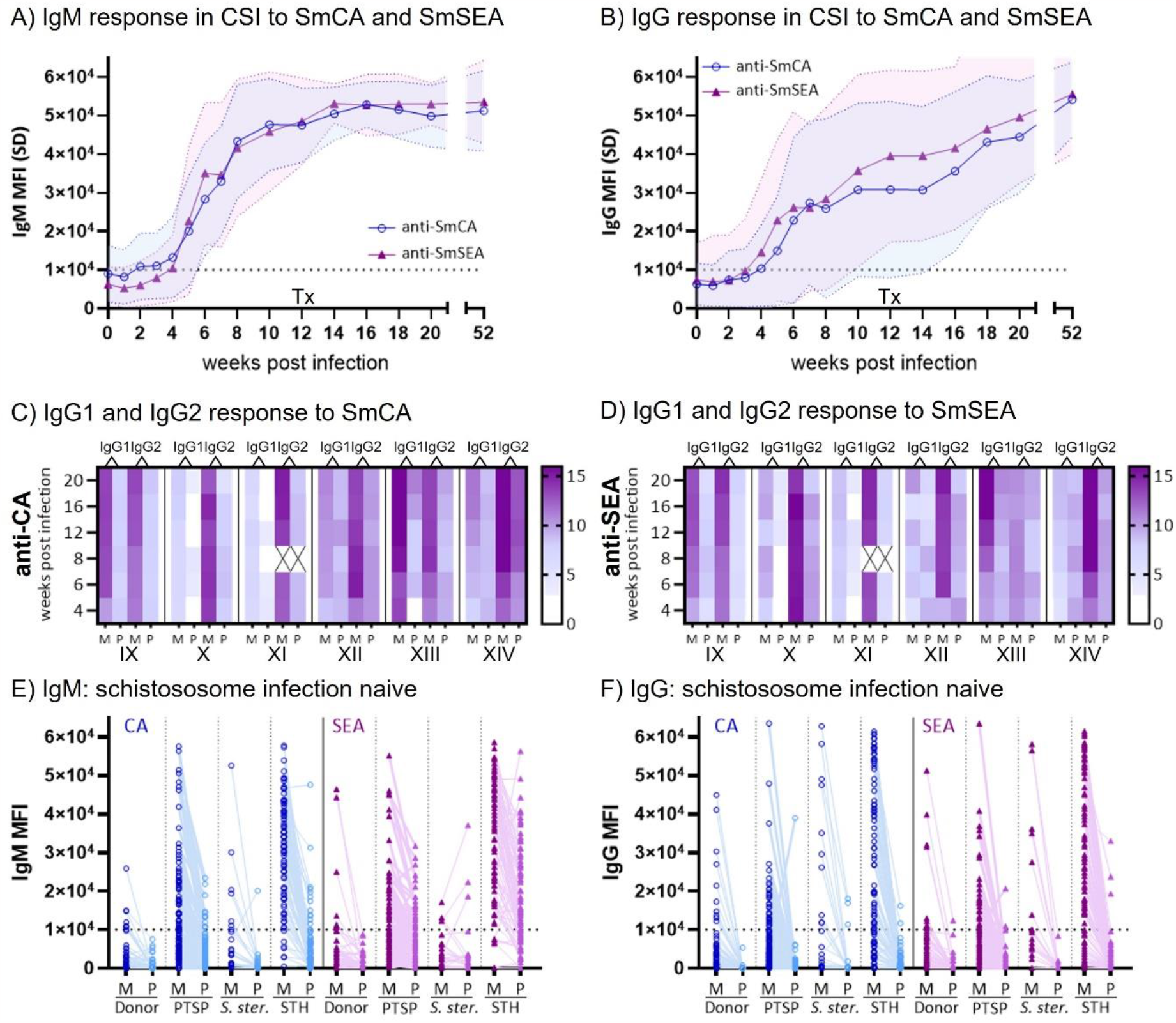
IgM and IgG from individuals with and without schistosome infection recognise glycan epitopes present in *S. mansoni* soluble cercarial (SmCA) and egg antigens (SmSEA) Average SmCA (circles) and SmSEA (triangles) specific A) IgM and B) IgG antibody levels elicited in response to controlled human schistosome infection (CSI) with male-only cercariae (dose 10/20/30 cercariae, n=17 individuals) over time. No parasite eggs were produced in this single-sex CSI model and hence all antibodies recognizing SmSEA are binding to shared, predominantly glycan, epitopes present in both SmCA and SmSEA. Tx indicates treatment with 40mg/kg praziquantel at twelve weeks post infection. Heatmaps showing levels (log2 MFI+1) of IgG1 and IgG2 specific for C) SmCA and D) SmSEA mock (M) and periodate treated (P) antigen for six individuals infected with 10 or 30 cercariae (10 cercs: IX, X, XI, 30 cercs: XII, XIII, XIV) at an early phase (weeks 4, 6, 8) and later phase (weeks 12, 16, 20) post infection. Note the highly similar pattern for SmCA and SmSEA, which is due to epitopes shared between SmCA and SmSEA (male only infection, no eggs present). No anti-SmCA/SmSEA IgG3 or IgG4 was detectable for IX, X, XI, not measured for XII, XIII, XIV. Any binding below log2 (MFI+1) = 8 is negligible. X = no data available. E) IgM and F) IgG antibodies recognising SmCA (blue circles) and SmSEA (purple triangles) are present in samples from Dutch blood donors (donor n=56), from Dutch pre-travel samples (PTSP n=131), from LUMC biobank samples with *S. stercoralis* infection (*S. ster*. n=25), and from samples with *Trichiura trichiura, Ascaris* sp., and/or hookworm infections from Flores Island, Indonesia which is also non-endemic for schistosomes (STH IgM n=87, IgG n=97). For each individual in each sample set paired data for mock treated (M, left) and periodate treated (P, right) soluble antigen is shown. A decrease in MFI for the vast majority of antibody binding in mock compared to periodate treated antigen illustrates, that many antibodies measured are recognising glycan elements present within the crude antigen mixes. Such shared glycan epitopes pose a challenge for specificity and hence accuracy in diagnostic immunoassays. See Suppl. Table 2 for specificity calculations based on the arbitrary cut-off (MFI=1×10^4^) shown as a dotted line on E) and F). All data shown as mean fluorescence intensity (MFI) measured on microarray. SD = standard deviation.

**Figure 2:**
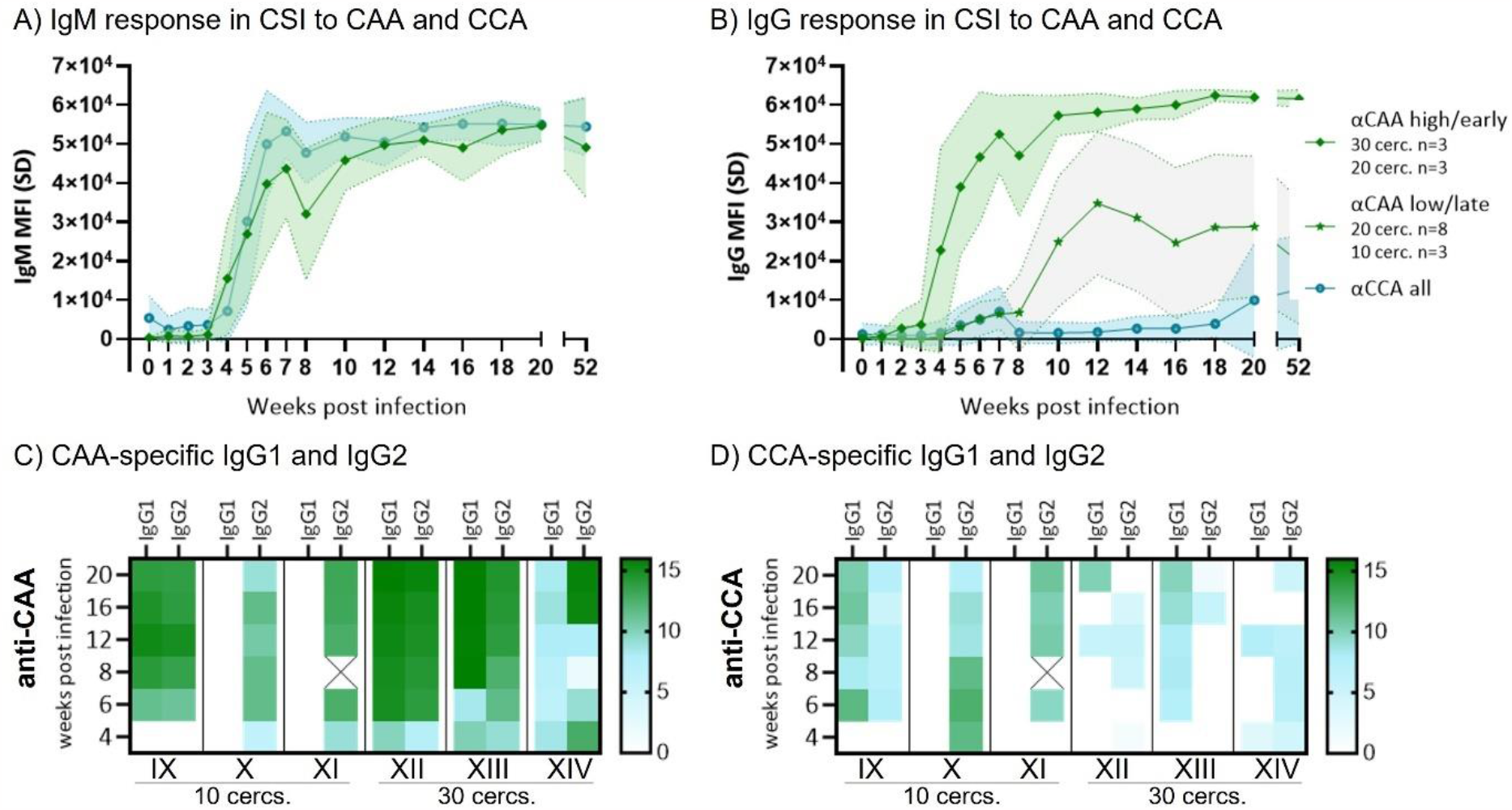
Schistosome circulating anodic and cathodic antigens are immunogenic with IgG response to CAA showing Infection dose dependency. Average anti-circulating anodic antigen (CAA, rhombes) and anti-circulating cathodic antigen (CCA, circles) specific A) IgM and B) IgG antibody levels induced in CSI with male-only cercariae (dose 10/20/30 cercariae, n=17 individuals) over time at baseline (0) and at one, two, three, four, five, six, seven, eight, ten, 12, 14, 16, 18, 20 and 52 weeks post-infection. For B) anti-CAA IgG responses are split in two groups depicting individuals responding early (≤ week 4, rhombes) and late (≥ week 7, stars). The early responder group consist of 30 (n=3) and 20 (n=3) exposure group individuals, whereas the late responder grouping consist of 20 (n=8) and 10 (n=3) exposure group individuals. Data shown as mean fluorescence intensity (MFI) with standard deviation (SD) measured on microarray. C) Heatmap showing anti-CAA and D) anti-CCA IgG1 and IgG2 levels (log2 MFI+1) for the same six individuals as Figure 1C,D (10 cercs: IX, X, XI, 30 cercs: XII, XIII, XIV) at an early phase (weeks 4, 6, 8) and later phase (weeks 12, 16, 20) post infection. No anti-CAA/CCA IgG3 or IgG4 was detectable for IX, X, XI, not measured for XII, XIII, XIV. Any binding below log2 (MFI+1) = 8 is negligible. X = no data available.

Focusing only on the CSI response results to SmCA and SmSEA, these appear to be good antigens as they follow a clear dose-response dynamic. However, when measuring anti-SmCA and anti-SmSEA antibodies in samples from schistosome infection naïve individuals, we found the specificity of these crude antigens to be as low as 8% (IgM) and 31-36% (IgG) in individuals infected with other helminths (Suppl. Table 2, Figure 1 E, F). Much of the reactivity to crude schistosome antigens in schistosome infection-negative individuals can be explained by antibodies recognising glycan epitopes present both in schistosomes (SmCA, SmSEA) and other organisms as seen by the consistent decrease in antibody binding to these antigens after periodate treatment(Figure 1 E,F).

**Table 2:** Specificity of CAA, CCA and crude schistosome antigens on microarray

| Specificity % *<br>(n true negatives/n) | CAA | CCA | SmCA | ShCA | SmSEA | ShSEA |
| --- | --- | --- | --- | --- | --- | --- |
| **IgM (all samples)** | 93,2 (278/298) | 84,6 (252/298) | 50,0 (148/298) | 29,9 (89/298) | 66,4 (198/298) | 63,4 (189/298) |
| Donor (no. inf) | 92,9 (52/56) | 98,1 (55/56) | 83,9 (47/56) | 58,9 (33/56) | 94,6 (53/56) | 94,6 (53/56) |
| PTSP (no. inf) | 96,9 (126/130) | 85,4 (111/130) | 58,5 (76/130) | 27,7 (36/130) | 80,0 (104/130) | 70,8 (92/130) |
| *S. ster.* | 100,0 (25/25) | 96,0 (24/25) | 64,0 (16/25) | 72,0 (18/25) | 96,0 (24/25) | 88,0 (22/25) |
| STH | 86,2 (75/87) | 71,3 (62/87) | 10,3 (9/87) | 2,3 (2/86) | 19,5 (17/87) | 25,3 (22/87) |
| **IgG (all samples)** | 97,4 (301/309) | 95,8 (296/309) | 73,1 (226/309) | 57,9 (179/309) | 71,8 (222/309) | 92,9 (287/309) |
| Donor (no. inf) | 100 (56/56) | 98,1 (55/56) | 89,3 (50/56) | 64,3 (36/56) | 91,1 (51/56) | 98,1 (55/56) |
| PTSP (no. inf) | 97,7 (128/131) | 97,7 (128/131) | 87,0 (114/131) | 73,3 (96/131) | 87,8 (115/131) | 99,2 (130/131) |
| *S. ster.* | 96 (24/25) | 92,0 (23/25) | 68,0 (17/25) | 68,0 (17/25) | 68,0 (17/25) | 96,0 (24/25) |
| STH | 95,9 (93/97) | 92,8 (90/97) | 46,4 (45/97) | 30,9 (30/97) | 40,2 (39/97) | 80,4 (78/97) |

\* Based on arbitrary cut-off at MFI=10000

### Anti-CAA antibodies show infection-dose dependent isotype switch in primary infection

The circulating schistosome antigens CAA and CCA showed promising sensitivity and specificity in the first down-selection (table 1). Both CAA and CCA are repetitive glycan structures. CAA is a glycoconjugate containing an antigenic O-linked carbohydrate structure consisting of N-acetyl galactosamine and glucuronic acid repeats [-6(GlcAβ1-3)GalNAcβ1-]n, whereas the CCA structure consist of repeating Lewis-X (LeX) motifs [-3Galβ1-4(Fucα1-3)GlcNAcβ1-]n (26, 28). Through assessment of full time-course CSI sample series (n=17), CAA- and CCA-specific IgM proved to be measurable already from three to four weeks post infection with comparable dynamics (Figure 2A). To CCA, very low IgG titers are elicited in response to infection (Figure 1B). This low antigenicity is possibly due to epitope similarity between LeX motifs of CCA and the LeX antigens present on a subset of human cells and serum glycoproteins (29). This is also consistent with published cross-sectional human data on anti-LeX antibody responses (11). In contrast, anti-CAA IgG responses are much stronger, and isotype switch present with an infection dose-dependent timing (Figure 1B).

Individuals exposed to 30 cercariae elicit anti-CAA IgG responses earlier and with higher titers compared to those exposed to only ten parasites. CSI participants exposed to twenty cercariae can be divided into two groups that follow the anti-CAA IgG response patterns of either the low dose (ten cercariae) or high dose (30 cercariae) groups. This corresponds to an early seroconversion of three to four weeks post infection in the high dose compared to ≥7 weeks post infection for the low dose. For the current CSI participants, the threshold CAA antigen quantity necessary to mount an early response hence falls between infection with ten and 20 cercariae. Similar to the observations for SmCA and SmSEA, the initial IgG subclass repertoire does not change for anti-CAA (or CCA) from early to later infection phase (Figure 2C,D) but variation is seen on inter-individual level.

Baseline background for both anti-CAA (and CCA) IgM and IgG is markedly lower than that for anti-SmCA and anti-SmSEA underpinning the high specificity of these antigens (Figure 1AB, 2AB).

### Host immune system is exposed to schistosome circulating antigens (CAA and CCA) early after infection

As specific antibody responses to CAA and CCA were measurable already within four weeks post-infection, parasite culture experiments were designed to confirm the presence of these antigens in early life stages. CAA and CCA are gut-associated antigens and likely present in low amounts in the non-functional cercarial gut primordium (30, 31). Increased expression of these antigens is expected when gut development progresses and the parasites start blood feeding, which also results in antigen regurgitation. To confirm the CAA/CCA presence and increased expression during the course of schistosomulae development, transformed cercariae were cultured in a 48-well plate and antigen content measured by UCP-LF CAA and CCA assays. CAA and CCA content was determined in individually collected schistosomulae day zero and eight (Figure 3 A,D). For CAA we observed a 4,8 fold increase in average CAA/parasite and equivalently a 6,0 fold increase for average CCA/parasite over the eight days. The increase in CAA and CCA produced by schistosomulae was confirmed by measurements done on total schistosomulae harvested from a multi-well experiment day one to seven as well as the corresponding culture supernatants (Figure 2B,C and E,F).The variation in excreted CAA/CCA levels between the five mini-cultures harvested each day may be explained by differences in viability and/or metabolic activity of the schistosomulae even if originating from the same pooled and transformed batch of cercariae. The antigens are measurable already 24-48 hours after culture set-up and reflect a source of antigenic stimuli released into the host circulatory system for uptake and processing.

**Figure 3:**
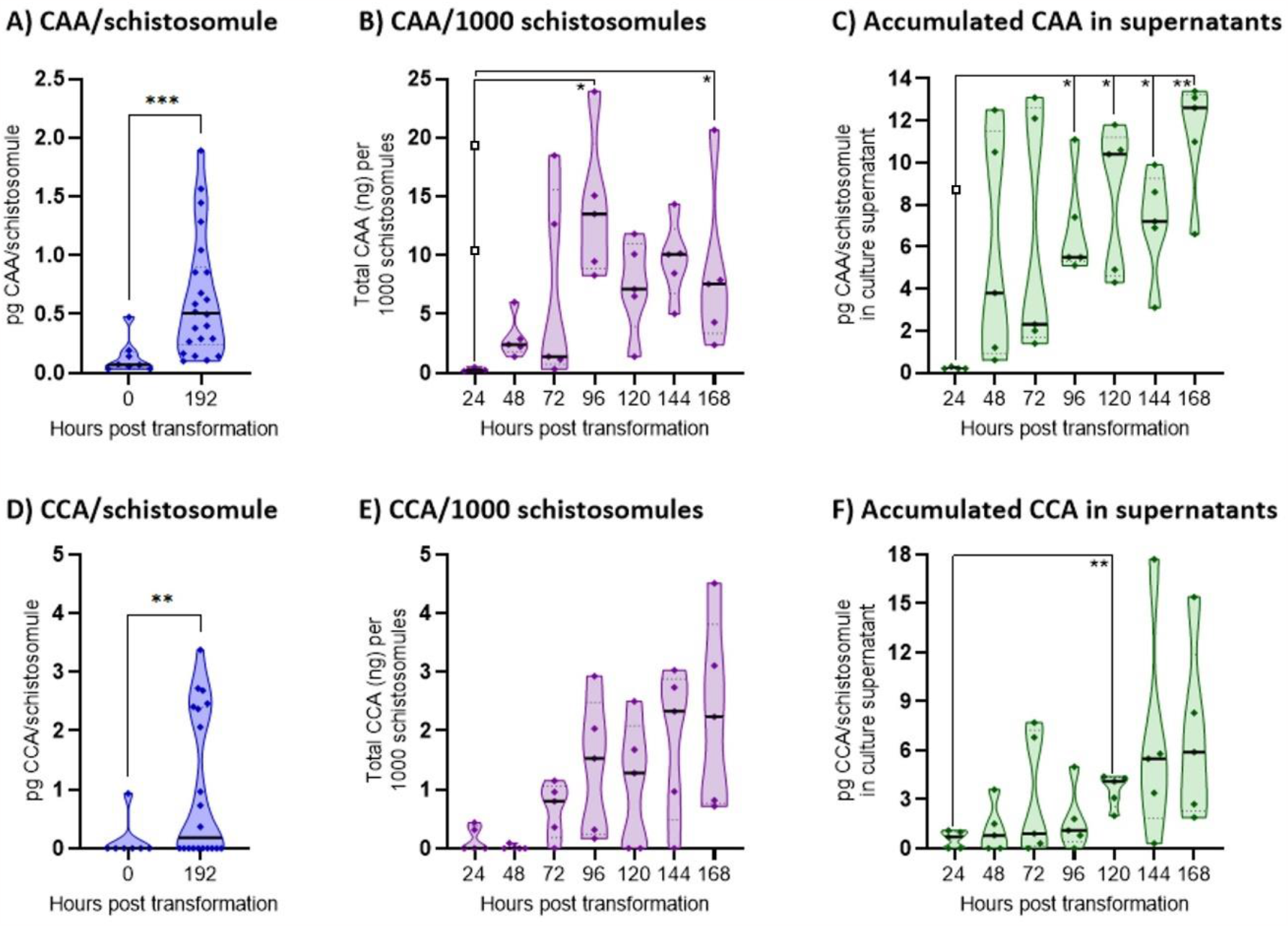
CAA and CCA amounts in schistosomula extracts and excretions. A-C) CAA amount measured by UCP-LF CAA wet assay and D-F) CCA amount measured by UCP-LF CCA wet assay shown as truncated violin plots with medians indicated by horizontal full line bars. A) CAA (pg) and D) CCA (pg) measured in schistosomulae individually collected just after transformation from cercariae and eight days of culturing. Individual pentaplicate wells were harvested from 48-well culture plate day one to seven and 10% volume with parasites counted thrice (parasites/well: range = 761-1294, mean = 968, median = 977). Transformation efficacy was ≥98%. B) CAA (ng) and E) CCA (ng) contained in the parasites and adjusted to amount/1000 schistosomulae at 24 hour intervals up to day seven (168 hours). C) accumulated CAA (pg)/schistosomula and F) CCA (pg)/schistosomulae excreted into culture supernatants adjusted to parasite count/well. Media controls were included in assays for all timepoints and for B, C) three datapoints marked with open squares at 24hrs are assay derived technical outliers. After day four, it was apparent that some parasites thrived and developed and others started dying in the culture wells, hence the measured CAA/CCA amounts per organisms from >96hrs is underestimated (B, C, E, F). Note that the technical sensitivity of the UCP-LF CCA assay (buffer format) is lower with a higher cut-off than the CAA assay, therefore the low range values cannot be directly compared between assays and hence the antigens.

### Evaluating specificity and sensitivity of anti-CAA and anti-CCA antibodies

To further evaluate specificity of CAA and CCA as antibody targets compared to crude schistosome antigen preparations, more samples from schistosomiasis naïve individuals were assessed: firstly, we analysed samples from individuals unlikely to have been exposed to or infected with helminths; Dutch blood donors and pre-travel study samples. Secondly, we analysed samples from individuals with soil-transmitted helminth infections. Schistosomiasis false positives for both IgM and IgG are much less frequent for CAA and CCA compared to crude antigen from both *S. mansoni* and *S. haematobium* (Figure 4, arbitrary cut-off). The superior specificity of CAA and CCA compared to crude SmCA and SmSEA is outlined in Table 2. IgG performs with ≥96% specificity for anti-CAA and ≥93% for anti-CCA across all sample sets, whereas the specificity falls a low as 31% for anti-ShCA and 40% for anti-SmSEA in the STH infected individuals. Specificity for IgM is slightly lower across sample sets with the lowest anti-CAA performance for STH samples at 86% and anti-CCA at only 71%. However, for crude antigens specificity in STH infected individuals is between 2-25% highlighting the gross cross-reactivity mediated by shared glycan epitopes. This low specificity render the crude antigen mixes useless in diagnostic context when measured on microarray.

**Figure 4:**
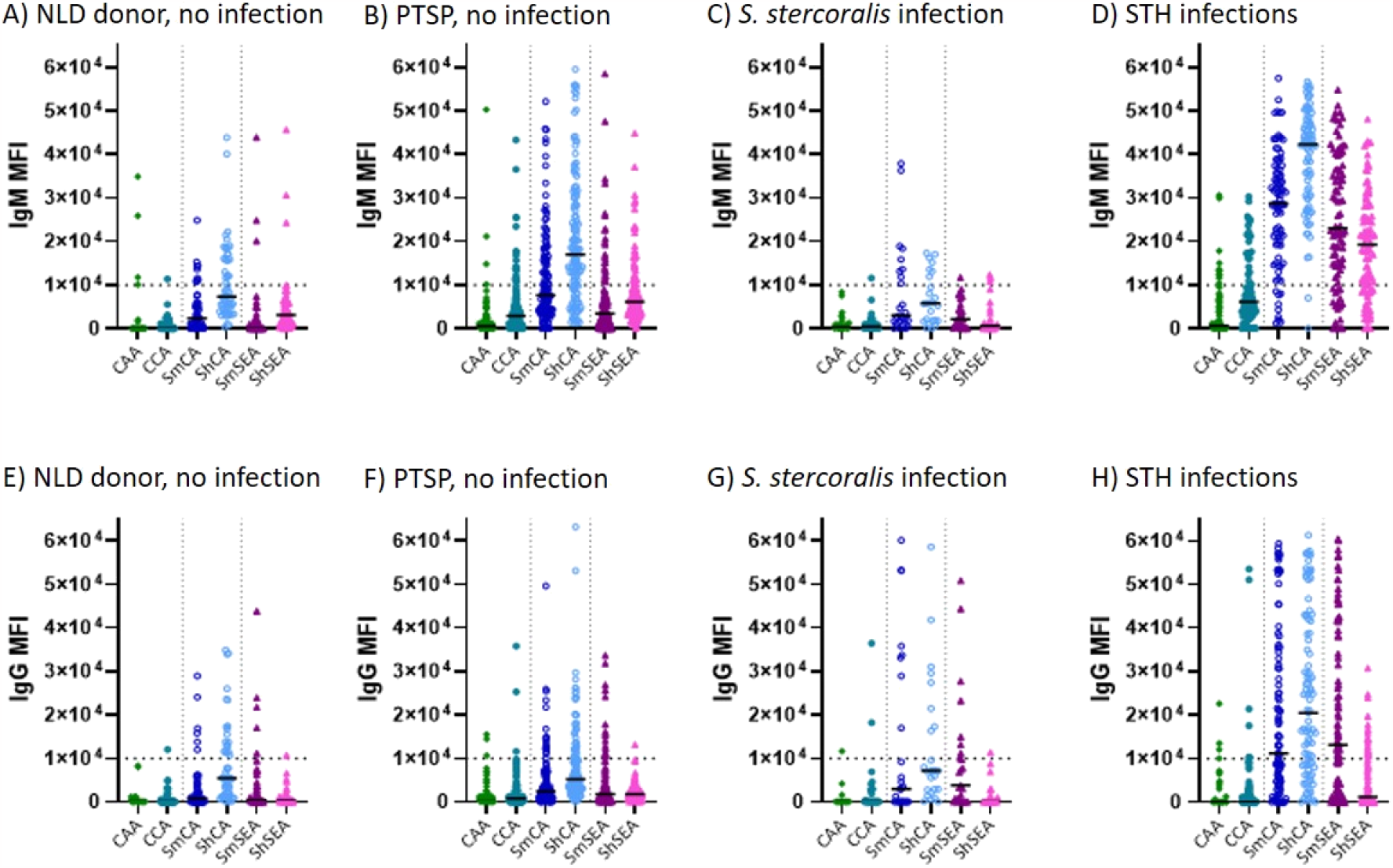
Antibodies recognizing circulating anodic antigen are highly specific. IgM (A-D) and IgG (E-H) responses to CAA, CCA, *S. mansoni* CA (SmCA), *S. haematobium* CA (ShCA), *S. mansoni* SEA (SmSEA), and *S. haematobium* SEA (ShSEA) for schistosomiasis infection naïve individuals are shown as dotblots for: Dutch blood donors (A and E, n=56 NLD donor)), Dutch pre-travel samples (B and F, n=IgM 130/IgG 131 PTSP), *S. stercoralis* infected LUMC biobank samples (C and H, n=25), and soil-transmitted helminth (*Trichuris trichiura, Ascaris* sp., and/or hookworm) infected individuals from Pulau Flores, Indonesia (D and H, n=IgM 87/IgG 97 STH). All data shown as mean fluorescence intensity (MFI) measured on microarray with medians indicated as horizontal bars. See table 2 for specificity calculations based on the arbitrary cut-off (MFI=10000) shown as a dotted lines on graphs (A-H).

Three sample sets from Europeans, that were infected during travel in schistosomiasis endemic areas, were used to further investigate sensitivity of anti-CAA and -CCA antibodies. High IgM and IgG antibody levels in six young Spanish travellers were seen from ≥44 days post infection (Figure 5A, B). All six individuals have high anti-CAA and anti-CCA IgM levels, whereas IgG is high for anti-CAA and less pronounced for anti-CCA consistent with CSI observations (Figure 2B). A similar pattern was observed in samples from a historical pre-post travel study that included nine schistosomiasis cases (Figure 5C). Importantly, anti-CAA IgG performed better than the SEA-ELISA and/or IFA (adult worm sections) used in the original study which consist of data from travel clinics (9/9 compared to 3/9). A third primary infection sample set consist of travellers exposed to *S. haematobium* clade parasites.

**Figure 5:**
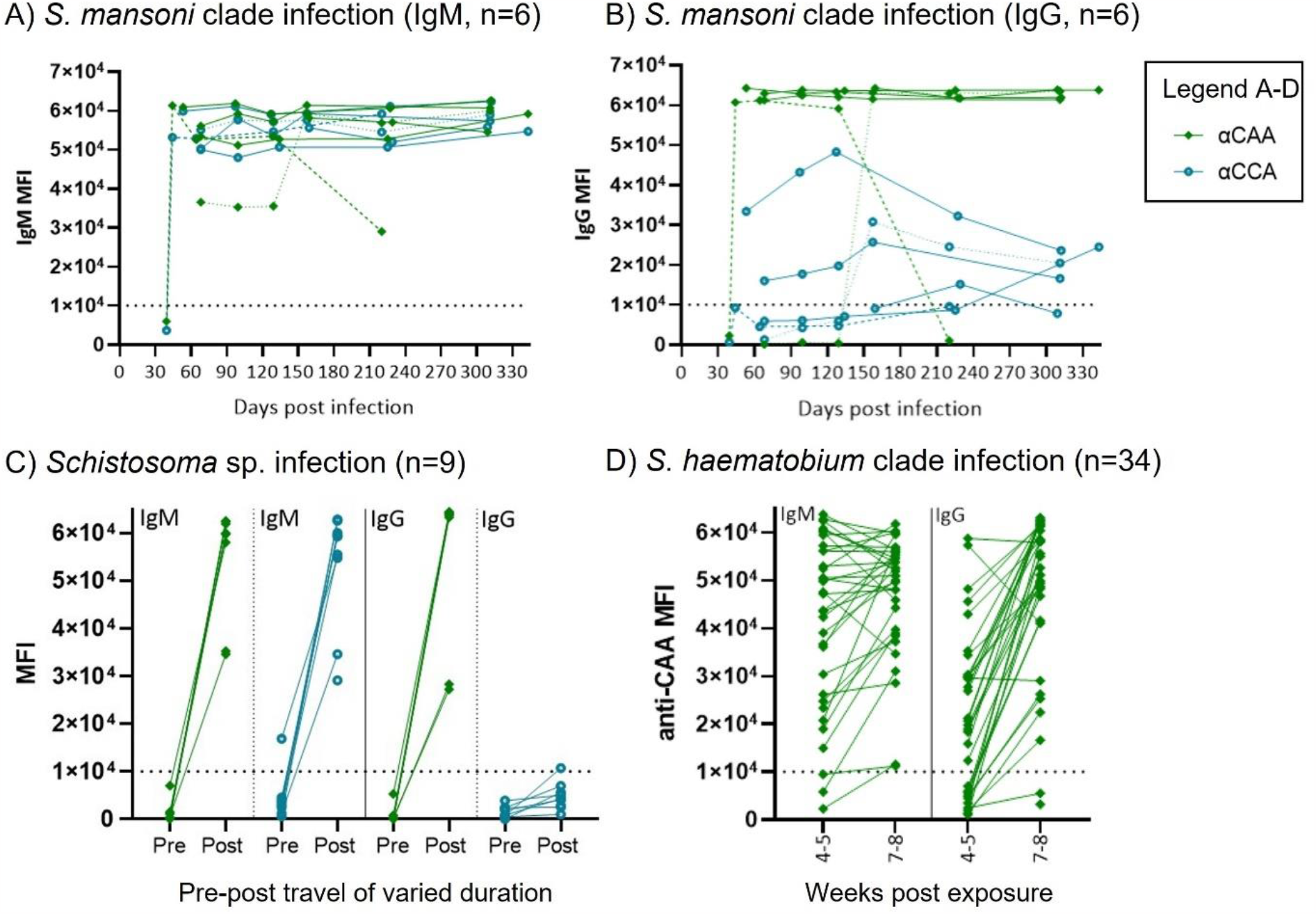
Anti-CAA and anti-CCA antibodies in primary infections in travellers. IgM (A) and IgG (B) responses to CAA and CCA in six young male Spanish travellers exposed to *S. mansoni* clade parasite in Chicamba Lake, Mozambique in 2019 through a single swimming activity (19). Individuals shown with dashed lines are paired for A and B to highlight time dynamics for individuals without consistently high antibody titer. These two individuals show differences in timing of isotype switch. One individual was the first to present with symptoms at the travel clinic at day 39 after infection (low antibodies) and with high IgM and IgG at 44 days post infection and declining antibody levels from ∼20 weeks post infection. The second has lower IgM from first measurement at ∼7 weeks up to ∼20 weeks post exposure, where isotype switch occurs and CAA specific IgG is induced. C) Anti-CAA and anti-CCA for paired pre- and post-travel samples from nine adult Dutch individuals all identified as schistosomiasis IgM positive but only three IgG positive in original study (18). These nine individuals were exposed to *Schistosoma* sp. through swimming in Lake Malawi and/or Lake Victoria at separate event(s) during travel of varying lengths and measured within three months after returning to The Netherlands. All post-travel samples from individuals with no schistosome exposure were confirmed to still be negative for anti-CAA and anti-CCA (PTSP pre-travel shown in Figure 4BF). D) shows anti-CAA responses for 34 Belgian individuals travelling as part of two separate groups of families but exposed at same location through swimming and rafting in uMhkunyane river, Northern Kwa-Zulu Natal, South Africa in 2016/17 to *S. haematobium* clade parasites (*S. haematobium x S. matheii* hybrid and/or co-infection(s)) within a two week interval (20, 21), no data for anti-CCA antibodies available for this sample set due to limited sample volume. Data shown as mean fluorescence intensity (MFI) measured on microarray.

Anti-CAA antibodies were measurable in 34/34 for IgM and 32/34 for IgG at seven to eight weeks post infection (Figure 5D). This finding is consistent with the cumulative positive pre-treatment serum CAA findings (33/34 positive) previously published (21). Merging the CSI and primary infection traveller sample sets gives an overall sensitivity for anti-CCA IgM of 100% (IgG not usable) and anti-CAA IgM 100%; IgG 97% (n=66, arbitrary cut-off MFI=10000, timing variable). These three primary infection traveller sample sets confirm the high sensitivity of anti-CAA and -CCA antibodies observed in CSI (Figure 2A,B).

## DISCUSSION

Through the microarray-based evaluation of a panel of representative schistosome antigens, CAA emerged as the best performing target with high sensitivity (IgM≥100%, IgG≥97%) and specificity (IgM≥93%, IgG≥97%) for serological detection of primary schistosomiasis infection. Interestingly, we observed an infection dose-dependent isotype switch in CSI time course samples. IgM responses to CAA were on average mounted already at three to four weeks post-infection irrespective of infection dose. However, isotype switch to IgG varied in an infection dose-dependent manner with a three to four week delay for individuals infected with a low dose (ten cercariae) compared to those with a higher (30 cercariae). This observation might be explained by differences in number of cercariae that successfully penetrate the skin and/or reach the lung combined with inter-individual immune response variability in the volunteers. Which isotype(s) would be optimal in terms of diagnostic value depends on the diagnostic assay platform and target product profile use-cases in future test developments. For example, IgM might be suitable in European travellers with low helminth infection frequency, whereas the higher specificity of IgG could be the better choice in a setting with frequent STH infection, as indicated by the Indonesian sample data. Our findings show that measuring antigen-specific total IgG is better than a single subclass, as not all individuals elicit the same subclass to CAA as observed in the CSI samples (IgG1 or IgG2 or IgG1/IgG2). IgG subclass response to glycan antigens may differ depending on epidemiological context. In this study, no IgG3 and IgG4 recognising glycan elements in crude SmCA or SmSEA was detectable in CSI samples, but both IgG3 and IgG4 specific for epitopes such as core-xylose elements present in crude antigen have been demonstrated in chronic infection samples (32, 33). Data on characteristics of antibody responses to schistosome infection such as isotype, IgG subclass, induction timing and persistence informs how and when serological tests can be used optimally within diagnostic logarithms for different epidemiological settings.

Limited data from different schistosomiasis endemic contexts support the antigenicity of the unique polysaccharides, CCA and CAA expressed in the schistosome gut. In concordance with our anti-CAA and anti-CCA observations, Nash *et al*. (1978) showed by IFA that individuals with acute infection presented with high antibody levels to gut associated polysaccharides, now known to include CCA and CAA (34). In contrast, chronic infection antibody levels were lower and disconnected from egg burden with increasing age (34, 35). Further studies must determine, under which infection pressures down-modulation and/or immune-complex formation may result in lower CAA-specific IgM and IgG measurements to define in which situations in endemic areas a test would be accurate (36, 37).

Irrespective of performance in long-term chronic infection, anti-CAA antibody detection can be a powerful surveillance tool in near- and post-elimination settings if applied in defined target groups such as occupational risk groups or children born after a reaching intervention goals. Detection of highly specific antibodies in people living in areas with risk of transmission (re-)emergence such as the Mediterranean would also be an attractive and cost-effective surveillance tool (38). For non-endemic travel medicine, our data from CSI and traveller samples indicate that development of an anti-CAA antibody-detection tool has potential to be highly accurate for both IgM and IgG detection. The high specificity of the existing CAA antigen detection assay supports the observation of superior specificity of anti-CAA antibodies as primary infection marker (17, 24). Anti-CAA antibody detection is expected to be pan-species specific, and should be evaluated further with well-characterised samples from people infected with other schistosome species such as *S. japonicum, S. haematobium, S. mekongi*, S. *intercalatum* and hybrid species/co-infections.

For upscaled and quality controlled production of a serological test, whether in finger-prick rapid diagnostic lateral flow test, ELISAs (direct/sandwich and or competition), immunoblot or bead/nanoparticle-based (multiplex) platform formats, the target much be produced in validated and feasible manners. This is achievable for relatively low through-put in house assays based on immunopurified native CAA such as formats used for non-endemic area travel medicine and research. Development of tests aimed at upscaled sero-surveillance or commercial formats will require establishment of chemical, enzyme-assisted or biotechnological synthesis of the catching antigen, CAA. While exogenous carbohydrate antigens are challenging to express in well-established systems, whether bacterial, yeast, baculoviral or mammalian cell line based, promising alternative platforms engineered for glycan antigen production are emerging. Successful glyco-engineering of the N-glycosylation machinery in *Nicotiana benthamiana* plants and production of glycoproteins with defined helminth-associated elements show that it is possible to recapitulate and produce complex glycan structures from schistosomes (39) when the corresponding components of biosynthetic machinery have been identified. In the case of CAA, which is a carbohydrate structure consisting of N-acetylgalactosamine (GalNAc) and glucuronic acid (GlcA) repeats (28) that would be the specific b6GalNAc-transferase and b3GlcA-transferase. Basing a test on carbohydrate rather than protein catching antigen encompasses the added value of better temperature stability and no vulnerability to protease degradation. The native CAA antigen has been found in more than 3000 year old mummies and is known to be highly stable at room temperature, refrigerated, frozen and in lyophilized form (40, 41). Such properties means less dependency of cold-chain transport and storage, which are factors that can have significant impact on cost and feasible real-life use in resource poor areas.

In conclusion, this study has identified CAA as antibody target with promising accuracy for primary schistosomiasis infection. Development of a highly sensitive and specific antibody detection assay will be a beneficial addition to the existing diagnostic tool repertoire for schistosomiasis. An anti-CAA antibody detection tool would have particular impact and use in traveller diagnostics and for surveillance in near- and post-elimination, eradication and emerging transmission zone settings.

## Supporting information

Suppl. methods and results

## Data Availability

All data in the present study will be made available online upon publication of the work in a peer-reviewed journal.

## ACKNOWLEDGEMENTS

First of all, we acknowledge and thank everyone participating in studies and the CSI trial for their invaluable contribution towards schistosome diagnostics development. We also thank all clinical and field work team members as well as Linda Wammes (LUMC) for their essential roles in making the sample material available. This work was supported by the Global Health Innovative Technology Fund (GHIT, RFP-TRP-2017-002), Leiden University Medical Center 2020 MSCA-IF Seal-of-Excellence programme, and the Human Infection Challenge Network for Vaccine Development (HIC-Vac, P68501, funded by the GCRF Networks in Vaccines Research and Development which was co-funded by the MRC and BBSRC. This UK funded award is part of the EDCTP2 programme supported by the European Union).

## Notes

### Competing Interest Statement

The authors have declared no competing interest.

### Author Declarations

All samples were fully anonymised (also to research group) and with consent for further serological work. CSI:The trial is registered at clinicaltrials.gov with identifier: NCT02755324 and ethical approval given by LUMC Institutional Medical Ethical Research Committee (Institutional Review Board P16.111). PTSP: The study protocol was approved by the Medical Ethics Committee at LUMC Spanish travellers: Hospital Clinic of Barcelona Institutional Review Board and Ethics Committee (HCB/2017/0612). Belgian travellers: Institutional Review Board of Institute of Tropical Medicine (ITM), Antwerpen Indonesian samples: Ethical Committee of the Medical Faculty, University of Indonesia (ethical clearance ref: 194/PT02.FK/Etik/2006) and filed by the Committee of Medical Ethics of the Leiden University Medical Center. The trial was registered as clinical trial (Ref: ISRCTN83830814).

