## Supplementary material for "Identification of CAA as highly specific and sensitive antibody target for acute schistosomiasis diagnostics": Suppl. methods and results

**SECTION 1: METHODS**

### Additional information on anonymised sample sets

#### Primary schistosome infection sample sets

**A)** Controlled human Schistosome Infection (CSI) serum samples from 17 healthy adult volunteers collected at baseline (bsl) and at one, two, three, four, five, six, seven, eight, ten, 12, 14, 16, 18, 20 and 52 weeks post-infection with male-only *S. mansoni* cercariae (no egg production). The original study was an open label dose-escalation clinical safety study carried out at Leiden University Medical Center, The Netherlands, where participants were exposed to respectively ten (n=3), 20 (n=11), and 30 (n=3) cercariae at week zero and treated with praziquantel at week 12 (40mg/kg body weight) as described in Langenberg *et al.* 2020 (1). The trial is registered at clinicaltrials.gov with identifier: NCT02755324 and ethical approval given by LUMC Institutional Medical Ethical Research Committee (Institutional Review Board P16.111).

**B)** Serum samples originally from the Post-Travel Screening of Parasites (PTSP) study published in Soonawala *et al.* 2014 (2) carried out in 2007-2009 with recruitment at travel clinics in Leiden and Wageningen, The Netherlands with specific written opt-in consent for use in other diagnostic research. Samples are from individuals travelling for minimum one month on the African continent and they were obtained pre-travel and 12 weeks after return to The Netherlands (n = 131 pre-travel, n=113 paired pre-post travel). The original study encompassed nine schistosome infection cases diagnosed post-travel by seroconversion (IFA and/or ELISA) all with histories of swimming in Lake Malawi and/or Lake Victoria.

**C)** *S. mansoni*-clade primary infection time course serum sample set from six young male Spanish adults exposed when swimming in Chicamba Lake, Mozambique on July 14$^{th}$ 2019. All individuals were egg negative and presented with acute schistosomiasis symptoms as described in Camprubí-Ferrer *et al.* 2021 (3). The participants were part of a larger cohort study on febrile travellers and approval was obtained through the Hospital Clinic of Barcelona Institutional Review Board and Ethics Committee (HCB/2017/0612).

**D)** Serum samples from *S. haematobium x S.matheii* primary exposure/infection in a cluster of Belgian tourists visiting South Africa exposed on two different occasions through swimming and rafting in uMhkunyane river (n=34, consisting of 8 families end of 2016/early 2017). All individuals were egg negative and 32/34 presented with acute schistosomiasis symptoms as published in Cnops *et al.* 2021, Hoekstra et al., 2021 (4, 5). Informed consent including storage and use for future schistomiasis diagnostic assays was given by all participants at the Institute of Tropical Medicine (ITM), Antwerpen, Belgium.

#### Schistosome infection-negative sample sets

**E)** Soil-transmitted helminth infection (STH) positive (*Ascaris* sp., *Trichuris trichiura*, *Necator americanus*, *Ancylostoma duodenale*, *Strongyloides stercoralis*) plasma samples from the Immunospin study as published in Wiria *et al.* 2010 with participants from Pulau Flores, Indonesia (n=97 from 2009 sample collection) (6). This island is not endemic for schistosomiasis and the likelihood of participants having a history of schistosome exposure is negligible. Clinical trial reference: ISRCTN83830814.

**F)** *Strongyloides stercoralis* infection positive sera (n=25) as diagnosed by ELISA and/or PCR were retrieved from a biobank based on the laboratory information management system (LIMS) of the clinical microbiology department of the LUMC. These 25 samples were handed to the research team

in a fully anonymised way. No schistosomiasis diagnostics was conducted at no further clinical information was given. Samples were only included if the patient had not specifically indicated that sample material could not be used for other purposes than diagnosis of disease, as regulated by law and stated in “Human tissue and medical research: code of conduct for responsible use” (2011).
G) Sera obtained via the Dutch blood donor bank, Sanquin, from healthy blood donors (n=56).

Supplementary table 1: Microarray target list
Down-selection arrays (8 samples gasket)

| Abbreviation | Structure/full name | Linker/protein carrier | Comment | Material origin (reference) |
|---|---|---|---|---|
| F | Fucα1- | 6-aminohexan-1-ol linker | synthetic | Harvey *et al.* (7) |
| FF | Fucα1-2Fucα1- | 6-aminohexan-1-ol linker | synthetic | Harvey *et al.* (7) |
| FFF | Fucα1-2Fucα1-2Fucα1- | 6-aminohexan-1-ol linker | synthetic | Harvey *et al.* (7) |
| FFFF | Fucα1-2Fucα1-2Fucα1-2Fucα1- | 6-aminohexan-1-ol linker | synthetic | Harvey *et al.* (7) |
| GalNAc | GalNAcβ1- | 6-aminohexan-1-ol linker | synthetic | Harvey MR (8) |
| FGalNAc | Fucα1-3GalNAcβ1- | 6-aminohexan-1-ol linker | synthetic | Harvey MR (8) |
| FFGalNAc | Fucα1-2Fucα1-3GalNAcβ1- | 6-aminohexan-1-ol linker | synthetic | Harvey MR (8) |
| Gn | GlcNAcβ1- | 6-aminohexan-1-ol linker | synthetic | Van Roon *et al.* (9) |
| F2Gn | Fucα1-2Fucα1-3GlcNAcβ1- | 6-aminohexan-1-ol linker | synthetic | Van Roon *et al.* (9) |
| F3Gn | Fucα1-2Fucα1-2Fucα1-3GlcNAcβ1- | 6-aminohexan-1-ol linker | synthetic | Van Roon *et al.* (9) |
| LDNaGal | GalNAc-β-(1- 4)-GlcNAcβ1-3Galα1- | 5-aminopenta-1-ol linker | synthetic | Ágoston *et al.* (10) |
| LDNFaGal | GalNAcβ1-4(Fucα1-3)GlcNAcβ1-3Galα1- | 5-aminopenta-1-ol linker | synthetic | Ágoston *et al.* (10) |
| FLDNFaGal | Fucα1-3GalNAcβ1-4(Fucα1-3)GlcNAcβ1-3Galα1- | 5-aminopenta-1-ol linker | synthetic | Ágoston *et al.* (10) |
| FLDNaGal | Fucα1-3GalNAcβ1-4GlcNAcβ1-3Galα1- | 5-aminopenta-1-ol linker | synthetic | Ágoston *et al.* (10) |
| LDNFaGal-BSA | GalNAcβ1-4(Fucα1-3)GlcNAcβ1-3Galα1- | bovine serum albumin | synthetic | Ágoston *et al.*,van Remoorte *et al.* (10, 11) |
| FLDNaGal-BSA | Fucα1-3GalNAcβ1-4GlcNAcβ1-3Galα1- | bovine serum albumin | synthetic | Ágoston *et al.*,van Remoorte *et al.* (10, 11) |
| aGal-BSA | Galα1-3Galβ1-4GlcNAcβ1- | bovine serum albumin | Commercial synthetic | Dextra Laboratories Ltd (UK) |
| LacNAc( | Galβ1-4GlcNAcβ1- | bovine serum albumin | synthetic | Van Roon *et al.* (12, 13) |
| LeX-BSA | Galβ1-4(Fucα1-3)GlcNAcβ1 | bovine serum albumin | synthetic | Van Roon *et al.* (12, 13) |
| 3`sialylLeX-HSA | NeuAcα2-3Galβ1-4(Fucα1-3)GlcNAcβ1 | bovine serum albumin | Commercial synthetic | Isosep (Tullinge, Sweden |
| di-LeX-BSA | Galβ1-4(Fucα1-3)GlcNAcβ1-3Galβ1-4(Fucα1-3)GlcNAcβ1- | bovine serum albumin | synthetic | Van Roon *et al.* (12, 13) |
| di-LeX-HSA | Galβ1-4(Fucα1-3)GlcNAcβ1-3Galβ1-4(Fucα1-3)GlcNAcβ1- | human serum albumin | Commercial synthetic | Isosep (Tullinge, Sweden) |
| LNFPIII-HSA | Galβ1-4(Fucα1-3)GlcNAcβ1-3Galβ1-4Glcβ1- | human serum albumin | Commercial synthetic | Isosep (Tullinge, Sweden) |
| LeA-HSA | Galβ1-3(Fucα1-4)GlcNAcβ1 | human serum albumin | Commercial synthetic | Isosep (Tullinge, Sweden) |
| CAA di-BSA | GlcAβ1-3GalNAcβ1- | bovine serum albumin | synthetic | Halkes *et al.*, Vermeer *et al.* (14, 15) |
| CAA tri-BSA | GalNAcβ1-6(GlcAβ1-3)GalNAcβ1- | bovine serum albumin | synthetic | Halkes *et al.*, Vermeer *et al.* (14, 15) |

| | | | | |
|---|---|---|---|---|
| CAA tetra-BSA | GlcAβ1-3GalNAcβ1-6(GlcAβ1-3)GalNAcβ1- | bovine serum albumin | synthetic | Halkes *et al.,* Vermeer *et al.* (14, 15) |
| CAA penta-BSA | GalNAcβ1-6(GlcAβ1-3)GalNAcβ1-6(GlcAβ1-3)GalNAcβ1- | bovine serum albumin | synthetic | Vermeer *et al.* (15) |
| CAA | [-6(GlcAβ1-3)GalNAcβ1-]n | none | *S. mansoni,* Immunopurified, capture antibody 51-4G5-A | van Dam *et al.* (16) |
| KLH | Hemocyanin | none | commercial, from *Megathura crenulata* | Sigma-Aldrich, H7017 |
| KLH- per | Hemocyanin sodium meta-periodate treated | none | commercial, from *Megathura crenulata* | Sigma-Aldrich, H7017 |
| SmAWA | adult worm antigen | none | mix of crude soluble antigens | *S. mansoni* LUMC life cycle* |
| SmAWA-mock | adult worm antigen mock treated | none | mix of crude soluble antigens | *S. mansoni* LUMC life cycle* |
| SmAWA- per | adult worm antigen sodium meta-periodate treated | none | mix of crude soluble antigens | *S. mansoni* LUMC life cycle* |
| SmCA | cercarial antigen | none | mix of crude soluble antigens | *S. mansoni* LUMC life cycle* |
| SmCA-mock | cercarial antigen mock treated | none | mix of crude soluble antigens | *S. mansoni* LUMC life cycle* |
| SmCA- per | cercarial antigen sodium meta-periodate treated | none | mix of crude soluble antigens | *S. mansoni* LUMC life cycle* |
| SmSEA | egg antigen | none | mix of crude soluble antigens | *S. mansoni* LUMC life cycle* |
| SmSEA-mock | egg antigen mock treated | none | mix of crude soluble antigens | *S. mansoni* LUMC life cycle* |
| SmSEA- per | egg antigen sodium meta-periodate treated | none | mix of crude soluble antigens | *S. mansoni* LUMC life cycle* |
| printbuffer | Proprietary | none | Commercial | Schott, Nexterion Spot 1066029 |

Reproduction and confirmation arrays (64-gasket): Targets of interest and controls from down-selection included (see Table 1) plus the targets below.

| | | | | |
|---|---|---|---|---|
| CCA* | [-3Galβ1-4(Fucα1-3)GlcNAcβ1-]n | none | *S. mansoni, ,* immunopurified, capture antibody 54-5C10-A | van Dam *et al.* (16) |
| ShCA | cercarial antigen | none | mix of crude soluble antigens | *S. haematobium* LUMC life cycle^ |
| ShSEA | egg antigen | none | mix of crude soluble antigens | *S. haematobium* LUMC life cycle^ |
| HSA | Human serum albumin | none | commercial | Sigma-Aldrich |

**Schistosoma mansoni* is the Puerto Rican strain and propagated at LUMC since 1955 (17).

^*Schistosoma haematobium* is from NAMRU-3, Cairo, Egypt propagated at LUMC since ~1993

**SECTION 2: RESULTS**

### Supplementary table 2: Shared glycan epitopes present in soluble crude SmCA and SmSEA decrease specificity

| ***IgM specificity*** | Donor* | PTSP* | *S. ster.** | STH* |
|---|---|---|---|---|
| CA-mock | 83,9 (47/56) | 57,7 (75/130) | 72,0 (18/25) | 8,0 (7/87) |
| CA-periodate | 100 (56/56) | 90,0 (117/130) | 96,0 (24/25) | 86,2 (75/87) |
| SEA-mock | 87,5 (49/56) | 63,8 (83/130) | 84,0 (21/25) | 8,0 (7/87) |
| SEA-periodate | 100 (56/56) | 80,0 (104/130) | 88,0 (22/25) | 36,4 (30/87) |
| ***IgG specificity*** | | | | |
| CA-mock | 82,1 (46/56) | 74,8 (98/131) | 60,0 (15/25) | 36,1 (35/97) |
| CA-periodate | 100 (56/56) | 99,2 (130/131) | 88,0 (22/25) | 97,2 (95/97) |
| SEA-mock | 82,1 (46/56) | 73,3 (96/131) | 72,0 (18/25) | 30,9 (30/97) |
| SEA-periodate | 98,2 (55/56) | 97,7 (128/131) | 100,0 (25/25) | 96,9 (94/97) |

*Specificity shown in percent and in brackets (n true negatives/n) based on defined arbitrary MFI=10000 cut-off (Figure 1E,F)

**METHOD REFERENCES**


1. Langenberg MCC, Hoogerwerf MA, Koopman JPR, Janse JJ, Kos-van Oosterhoud J, Feijt C, et al. A controlled human *Schistosoma mansoni* infection model to advance novel drugs, vaccines and diagnostics. Nature medicine. 2020;26(3):326-32.
2. Soonawala D, van Lieshout L, den Boer MAM, Claas ECJ, Verweij JJ, Godkewitsch A, et al. Post-travel screening of asymptomatic long-term travelers to the tropics for intestinal parasites using molecular diagnostics. The American journal of tropical medicine and hygiene. 2014;90(5):835-9.
3. Camprubí-Ferrer D, Romero L, Van Esbroeck M, Wammes LJ, Almuedo-Riera A, Rodriguez-Valero N, et al. Improving the diagnosis and management of acute schistosomiasis with antibody, antigen and molecular techniques: lessons from a cluster of six travellers. Journal of Travel Medicine. 2021;28(6).
4. Cnops L, Huyse T, Maniewski U, Soentjens P, Bottieau E, Van Esbroeck M, et al. Acute Schistosomiasis With a *Schistosoma mattheei × Schistosoma haematobium* Hybrid Species in a Cluster of 34 Travelers Infected in South Africa. Clinical Infectious Diseases. 2021;72(10):1693-8.
5. Hoekstra PT, van Esbroeck M, de Dood CJ, Corstjens PLAM, Cnops L, van Zeijl-van der Ham CJG, et al. Early diagnosis and follow-up of acute schistosomiasis in a cluster of infected Belgian travellers by detection of antibodies and circulating anodic antigen (CAA): A diagnostic evaluation study. Travel Medicine and Infectious Disease. 2021;41:102053.
6. Wiria AE, Prasetyani MA, Hamid F, Wammes LJ, Lell B, Ariawan I, et al. Does treatment of intestinal helminth infections influence malaria? Background and methodology of a longitudinal study of clinical, parasitological and immunological parameters in Nangapanda, Flores, Indonesia (ImmunoSPIN Study). BMC infectious diseases. 2010;10(1):77.
7. Harvey MR, Chiodo F, Noest W, Hokke CH, van der Marel GA, Codée JDC. Synthesis and Antibody Binding Studies of Schistosome-Derived Oligo-α-(1-2)-l-Fucosides. Molecules. 2021;26(8):2246.
8. Harvey MR. Synthesis and application of glycans unique to *S. mansoni*: Leiden University; 2020.
9. van Roon A-MM, Aguilera B, Cuenca F, van Remoortere A, van der Marel GA, Deelder AM, et al. Synthesis and antibody-binding studies of a series of parasite fuco-oligosaccharides. Bioorganic & Medicinal Chemistry. 2005;13(10):3553-64.
10. Ágoston K, Kerékgyártó J, Hajkó J, Batta G, Lefeber DJ, Kamerling JP, et al. Synthesis of Fragments of the Glycocalyx Glycan of the Parasite Schistosoma mansoni. Chemistry – A European Journal. 2002;8(1):151-61.
11. van Remoortere A, Vermeer HJ, van Roon AM, Langermans JA, Thomas AW, Wilson RA, et al. Dominant antibody responses to Fucalpha1-3GalNAc and Fucalpha1-2Fucalpha1-3GlcNAc containing carbohydrate epitopes in Pan troglodytes vaccinated and infected with *Schistosoma mansoni*. Experimental parasitology. 2003;105(3-4):219-25.
12. van Roon AM, Pannu NS, Hokke CH, Deelder AM, Abrahams JP. Crystallization and preliminary X-ray analysis of an anti-LewisX Fab fragment with and without its LewisX antigen. Acta Crystallogr D Biol Crystallogr. 2003;59(Pt 7):1306-9.
13. Roon A-Mv. *Schistosoma mansoni*: structural and biophysical aspects of Lewis X-antibody interactions 2005.
14. Halkes KM, Vermeer HJ, Slaghek TM, van Hooft PA, Loof A, Kamerling JP, et al. Preparation of spacer-containing di-, tri-, and tetrasaccharide fragments of the circulating anodic antigen of *Schistosoma mansoni* for diagnostic purposes. Carbohydr Res. 1998;309(2):175-88.
15. Vermeer HJ, Halkes KM, van Kuik JA, Kamerling JP, Vliegenthart JFG. Synthesis and conjugation of oligosaccharide fragments related to the immunologically reactive part of the circulating anodic antigen of the parasite *Schistosoma mansoni*. Journal of the Chemical Society, Perkin Transactions 1. 2000(14):2249-63.

16. van Dam GJ, Seino J, Rotmans JP, Daha MR, Deelder AM. *Schistosoma mansoni* circulating anodic antigen but not circulating cathodic antigen interacts with complement component C1q. European journal of immunology. 1993;23(11):2807-12.
17. Janse JJ, Langenberg MCC, Kos-Van Oosterhoud J, Ozir-Fazalalikhan A, Brienen EAT, Winkel BMF, et al. Establishing the Production of Male *Schistosoma mansoni* Cercariae for a Controlled Human Infection Model. The Journal of infectious diseases. 2018;218(7):1142-6.
